# Intentions and Willingness to receive Covid-19 vaccine among teaching and non-teaching staff in selected higher institutions of learning in Kampala and Wakiso districts, Central - Uganda

**DOI:** 10.1101/2023.05.16.23290042

**Authors:** Patrick Madrama Lulu, Fiona Atim, Kareodu Ronald, Fredrick Lugaro Wakula, John Charles Okiria

## Abstract

**Background:** COVID-19 was first discovered by WHO, 2019 in Wuhan, later spread to different parts of the world with thousands of deaths. COVID-19 vaccine was very important in reducing severity of the infection. Willingness to be vaccinated considerably varied according to regions. We assessed factors influencing intentions and willingness to receive covid-19 vaccines among teaching and non-teaching staff in higher institutions of learning.

**Methods:** A descriptive cross-sectional study design was adopted employing quantitative data collection and analysis approaches. Data was collected electronically using different online sources including emails, social media, popular media platforms and websites. 363 sample determined using Kish & Leslie (1965) of simple random sampling for single proportions. Univariate, bivariate and multivariate analysis done using SPP (23.0).

**Results:** 130(35.8%) intentions and willingness to receive COVID-19 vaccine. Influencing factors were; age between 30-39 (aOR=15491.54,95% CI=359-667551.9, P=0.000**, 40-49 (aOR=931, 95% CI=25.6-33816.4, p=0.000**), gender (aOR=4.66, 95% CI=2.06-1056, p=0.000*), staff category (aOR=7.71, 95% CI=4.34-14.71,p=0.000**), ownership (aOR=0.08, 95% CI=0.032-0.206, p=0.000**), being under health insurance (aOR=200.62, 95% CI=29.6-1359.18, p=0.000*, residency (aOR=19.88, 95% CI=4.63-85.26, p=0.000*), knowing where to get the vaccine (aOR=121.15, 95% CI=161.1-910.3, p=0.000**), COVID-19 vaccine important for prevention of the infection (aOR=19.73, 95% CI=2.27-171.52, p=0.007*), minor side effects of COVID-19 vaccines manageable (aOR=002, 95% CI= 0.00-0.15, p=0.000**), take painkillers if developed side effects (aOR=8.67, 95% CI=4.87-15.43, p=0.000*), responsibility to protect others by getting vaccinated (aOR=0.36, 95% CI=0.21-0.62, p=0.000**), concerned about getting infected with COVID-19 from the vaccine (aOR=0.002, 95% CI=0.00-0.03, p=0.000*).

**Conclusion:** Intentions and willingness to receive COVID-19 vaccine was low. This was attributed to age, gender, staff category, ownership, knowledge, attitude, where to get the vaccine, trust, vaccine efficacy, concern about getting infected after receiving the vaccine. MoH/MoE&S and development partners should emphasize improving attitude and knowledge of teaching and non-teaching staff in higher institutions to increase willingness.

## Background

COVID-19 is a disease caused by SARS-CoV-2, it was first discovered by the WHO on 31 December 2019 in Wuhan, the People’s Republic of China and later spread to different parts of the world [1], with thousands of cases and death that were being recorded almost every day [2], WHO recommended that, as people continued to observe public health standard operative procedures and IPC measures in communities and different settings including higher institutions of learning, vaccination of persons would be a very important venture to undertake [3]. Intentions are efforts or behaviour teachers are willing to exert to receive COVID-19 vaccines and they assume behaviours that reflect motivation for action [4]. Willingness reflects the openness of individuals to the opportunity of COVID-19 vaccination, involving some pre-contemplation on the implications of one’s behaviours [5].

Globally, by February 2021, total cumulative cases of the disease were 105,658,476, with 2,309,370 deaths and addition cases 14,166,078 and 329,863 additional deaths [6], as of April 2021, total addition 19,378,997 cases of COVID-19 and 343,631 deaths were confirmed [7]. By November 2020 the high-income countries had made premarket purchase commitments to buy over half of all pre-sold doses of COVID-19 vaccine, with some countries buying more doses than would be necessary to vaccinate their entire populations [8, 9]. Vaccines that were granted Emergency Use Listing by WHO included, AstraZeneca/Oxford, Johnson and Johnson, Moderna, Pfizer/Biotech, Sinopharm, Sinovac [10]. 26.1% of the general population had received at least one dose of a COVID-19 vaccine, 3.61 billion doses had been administered globally, and 30.46 million are now administered each day and only 1% of people in low-income countries had received at least one dose [11]. Vaccines have been a successful measure of disease prevention for decades [12]. However, vaccine hesitancy and refusal are significant concerns globally, prompting the World Health Organization (WHO) to declare this uncertainty among the top 10 health threats in 2019 [13]. A global survey conducted in 2020 on a potential acceptance of COVID-19 vaccine in 19 countries revealed that, 71.5% of participants would likely take the vaccine if it was proven safe and effective and the acceptance rates varied between countries ranged from 90% (in China) to 55% (in Russia), generally, acceptance of COVID-19 vaccine in Asian countries exceeded 80% [14].

In the United States, COVID-19 vaccine acceptance was at 67% among the general population [15]. In Europe, 73.9% were willing to get vaccinated against COVID-19 if available. The willingness ranged from 62% in France to about 80% in Denmark and UK. The willingness to be vaccinated considerably varied across gender [16]. In low developed countries like the Middle East, Russia, several European countries and Africa, COVID-19 vaccine acceptance were low [17]. In Jordan, COVID-19 vaccine acceptance was 37.4%, a need for public health authorities to work towards improving vaccine acceptance and reduce hesitancy [18].

In Sub-Saharan Africa, there is generally low acceptance of COVID-19 vaccines [19], with only less than 2% of global COVID-19 administered in Africa and 10 countries receiving 93% of the given doses [20]. Such variation may result in a difference in vaccine coverage and delay global control of the pandemic. As of 16^th^ July 2021, 169,074 cases of COVID-19 with 2,126 deaths were confirmed and a total of 3,938,945 vaccine doses have been administered in Nigeria, 277,443 cases confirmed with 4,350 deaths, and 2,062,456 vaccines administered in Ethiopia, 98,114 cases with 806 deaths and 1,265,306 vaccines administered in Ghana, 2,253,240 cases with 65,972 deaths and 4,236,718 vaccines administered in South Africa, 191,020 cases with 3,746 deaths and 1,477,916 vaccines administered in Kenya, 51,625 cases with 616 deaths, 398,096 vaccine administered in Rwanda [2].

Forty-three (43) African countries have so far joined the COVAX facility, which aims to ensure equitable access to safe and effective COVID-19 vaccines globally, the aim is to vaccinate at least 20% of the population by providing up to 600 million doses by the end of 2021, with the first phase of 90 million doses to support African countries immunize 3% of the population that are most in need of protection, including health workers and other vulnerable groups, in the first half of 2021 [21]. The vaccines pillar of the World Health Organization’s Access to COVID-19 Tools (ACT) Accelerator, the COVAX Facility seeks to ensure a more equitable distribution of covid-19 vaccines, regardless of income levels [22].

In Uganda, from the 3^rd^ of January 2020 to 16 July 2021, there have been 89,080 confirmed cases of COVID-19 with 2,249 deaths and 1,058,084 vaccine doses have been administered [2], with a low COVID-19 vaccine acceptance level in western Uganda among the general population [23] and high level of acceptance among health workers in Uganda [24]. There is scanty and limited data on intentions and willingness to receive the COVID-19 vaccine among teaching and non-teaching staff in higher Institutions of learning in Uganda. However, there has been an emergency COVID-19 vaccine withdrawal from some districts in Uganda where the said vaccines were not being well consumed by the target beneficiaries including teachers so that those vaccines could be used in Kampala and Wakiso which had the highest number of people infected with COVID-19 in Uganda [25].

## Materials and Methods

### Study Population and Settings

This cross-sectional study on intentions and willingness to receive COVID-19 vaccines among teaching and non-teaching staff in selected higher institutes of learning was conducted online through anonymous web-based survey in Wakiso and Kampala districts in Central Uganda between 10^th^ September to 30^th^ November 2021.

The study population is the subset of the target population for the study [26]. Our study population included all teaching and non-teaching staff in selected higher institutions in Kampala and Wakiso districts. This study population was selected because vaccinating teachers and other professionals working in these education institutions ensures continuity of teaching in-person, reducing teachers’ likelihood of infection, improves student safety; and increasing confidence in parents that education institutions are safe places to be in [27, 28] and there is very scares and limited data in Uganda on the intention and willingness of teaching and non-teaching staff in higher institutions of learning to receive COVID-19 vaccine. Our inclusion criteria required that, for a teaching and non-teaching staff to participate in the study, they must have been working in any of selected higher institution of learning for at least six (6) months and should have not yet received COVID- 19 vaccine so as to get authentic information.

### Sampling technique and Sampling Procedure

The researchers employed simple random sampling technique to select institutions and participants who fulfilled the inclusion criteria. There were 23 list of approved institutions of higher learning in Kampala and Wakiso districts as obtained online from the website of National Council for higher education in Uganda. One government higher education institution was randomly, one private institution selected from Wakiso and six from Kampala. The reason why six institutions were selected from Kampala is because Kampala has the highest number of institutions compared to Wakiso.

At the beginning of the survey for the participants selection, there was a detailed written informed consent section which explain the intent, inclusion and exclusion criteria, types of questions that would be asked, the anonymity and voluntary nature of the study. The participants were self- interviewed using electronic questionnaire and the setup was in such a way that, the questionnaire was only able to display after the respondent first read, understood and consented to the written electronic informed consent. The questionnaire was developed in English and circulated through emails, social media, popular media platforms and websites of the different institutions under study, no sensitive and personal information was collected and the authors had no access to any information that could identify participants during or after data collection.

### Study Variables

The dependent variable, Intentions and willingness to receive COVID-19 vaccine was measured using two items (Whether respondent had intentions and willingness to receive COVID-19 vaccine and when the respondent plans to receive the vaccine), the independent variables included social economic factors, knowledge, attitude and health system factors. We used 8 items (ever heard about COVID-19 vaccine, know where to get COVID-19 vaccine, knows that COVID-19 vaccine is important for prevention of COVID-19, do COVID-19 vaccine help develop herd immunity in the community? COVID-19 vaccine is safe and effective in ending the pandemic, COVID-19 vaccine prevent people from getting seriously ill, minor side effects of COVID-19 vaccine are manageable, taking painkillers like paracetamol before receiving COVID-19 vaccine to prevent side effects is not recommended, you may take painkillers if you have developed side effects such as pain, fever and headache after vaccination, even after vaccination keep taking precautions) to measure knowledge, 14 items were used to measure attitude (teachers should be prioritized for the COVID-19 vaccine, there is no chance of me becoming infected with COVID-19 because I follow the SOPS, At the risk of severe COVID-19, Complication on a scale of 1-100, what do you think is your level of risk to be infected with COVID-19, trust information of authorities on COVID19 vaccine, afraid about adverse side effects of the vaccine, it is my responsibility to protect others by getting vaccinated, I have concerns about the safety of the Vaccine, the vaccine was likely developed too first, I do not trust the development process of COVID-19 vaccine, I am not at risk of getting seriously ill of COVID-19 complications, concerned about getting infected with COVID-19 from the vaccine, education will not go back to normal until most teachers are vaccinated, I trust the efficacy of the COVID-19 Vaccine and 5 items were used to measure health system factors (simple and clear message on Covid-19 vaccine designed by the Ministry of Health, waiting time at the place to receive the Covid-19 vaccine is too long, the waiting time between the first and the second dose is too long, Ministry of Health is clear about the efficacy of the Covid- 19 vaccine, Health workers have confidence in the Covid-19 vaccine).

### Study Design

In this study we used a cross-sectional study design and employed quantitative data collection method and analysis. Data was collected electronically using different online sources including emails, social media, popular media platforms and websites. One page recruitment form was developed, including an online questionnaire link containing information about objectives, the importance of the study and ethical considerations. Online data collection method was prepared because this was done during the second COVID-19 lock down in Uganda.

363 sample determined using Kish & Leslie (1965) of simple random sampling for single proportions as explained in sample size calculation for medical studies [29]. The proportion of 70.2% willingness to receive COVID-19 vaccine among health workers in Uganda [24], represented “**p**” in the formula to determine the required sample size for teaching and non-teaching staff in higher institutions of learning in Uganda.

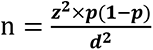

**n** refers to the sample size required, (**Z1-α/2)^2^** = Confidence level of 95% (Standard value of 1.96), **p** refers to the expected proportion of willingness and intention to receive COVID-19 vaccine among teaching and non-teaching staff members in higher institutions of learning in Uganda which is 70.2% =0.702, **q**= proportion of teachers and non-teaching staff in higher institutions of learning not willing to receive COVID-19 vaccine: 1-0.702 = 0.298, **d** refers to acceptable margin of error = 0.05. A simple random sampling technique was used, univariate, bivariate and multivariate analysis were done using SPSS (23.0).

### Data Analysis

We analysed the data using Microsoft Excell 2013 and SPSS version 23.0 at 95% CI. For consistency and completeness purpose, all data collected went undergo double-checking by the researchers. To determine intentions and willingness to receive and factors influencing intentions and willingness to receive COVID-19 vaccine, we did bivariate and multivariate logistic regression analysis for significant variables, results were expressed in form of unadjusted odds ratios (ours) and adjusted odds ratios (aOR) as required. Logistic regression was done to obtain the strength of association between independent and dependent variables and items that were statistically significant were analysed using multivariate logistic regression to obtain adjusted conclusions for the study findings.

### Ethical Considerations

We conducted this study in accordance with the Helsinki declaration guideline, ethical approval was granted by Clarke International University Research Ethics Committee (CLARKE-2021-141). After the ethical approval, administrative clearance was done by the institutions that were considered for the study and written online informed consent was obtained from study participants.

## Results

### Demographic Characteristics of respondents

In this study, the majority 160(44.1%) of the respondents were between 40-49 years of age, with the least 26(7.2%) being between 20-29 years. Male respondents were the majority 259(71.3%), the females accounted for 104(28.7%). In terms of marital status, the majority 218(60.1%) were single and 145(39.95%) were married. The majority 160(44.1%) were Pentecostals, followed by Roman Catholics 117(32.2%) and the least 1(0.3) being SDA.

In line with staff category, the majority 262(72.2%) were teaching staff, while 101(27.8%) were non-teaching staff. Respondents having postgraduate papers (Postgraduate diploma or masters) were the majority 259(71.3%) and the least 50(13.8%) had only bachelor’s degree. In line with terms of employment, the majority 229(63.1%) were in full time employment, while 134(36.9%) in part time (**See Table 1**).

**Table 1:**
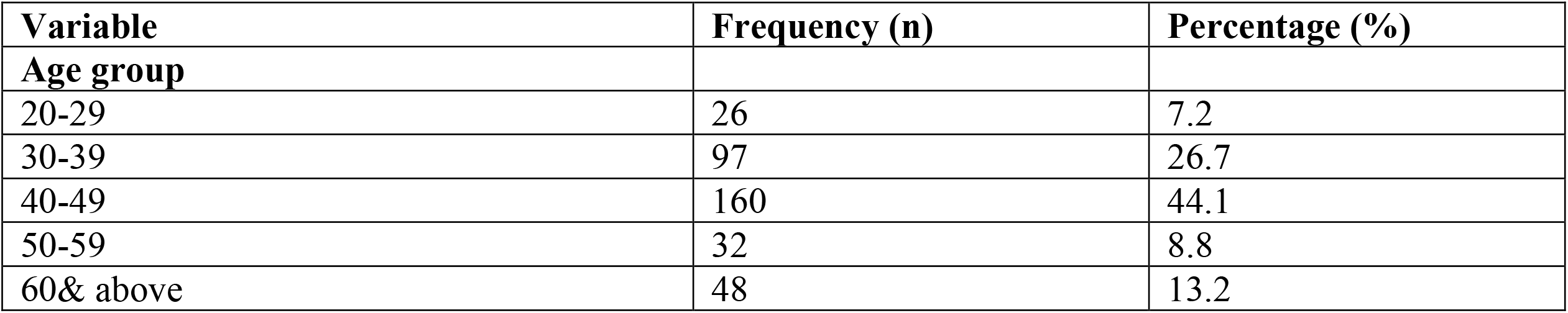
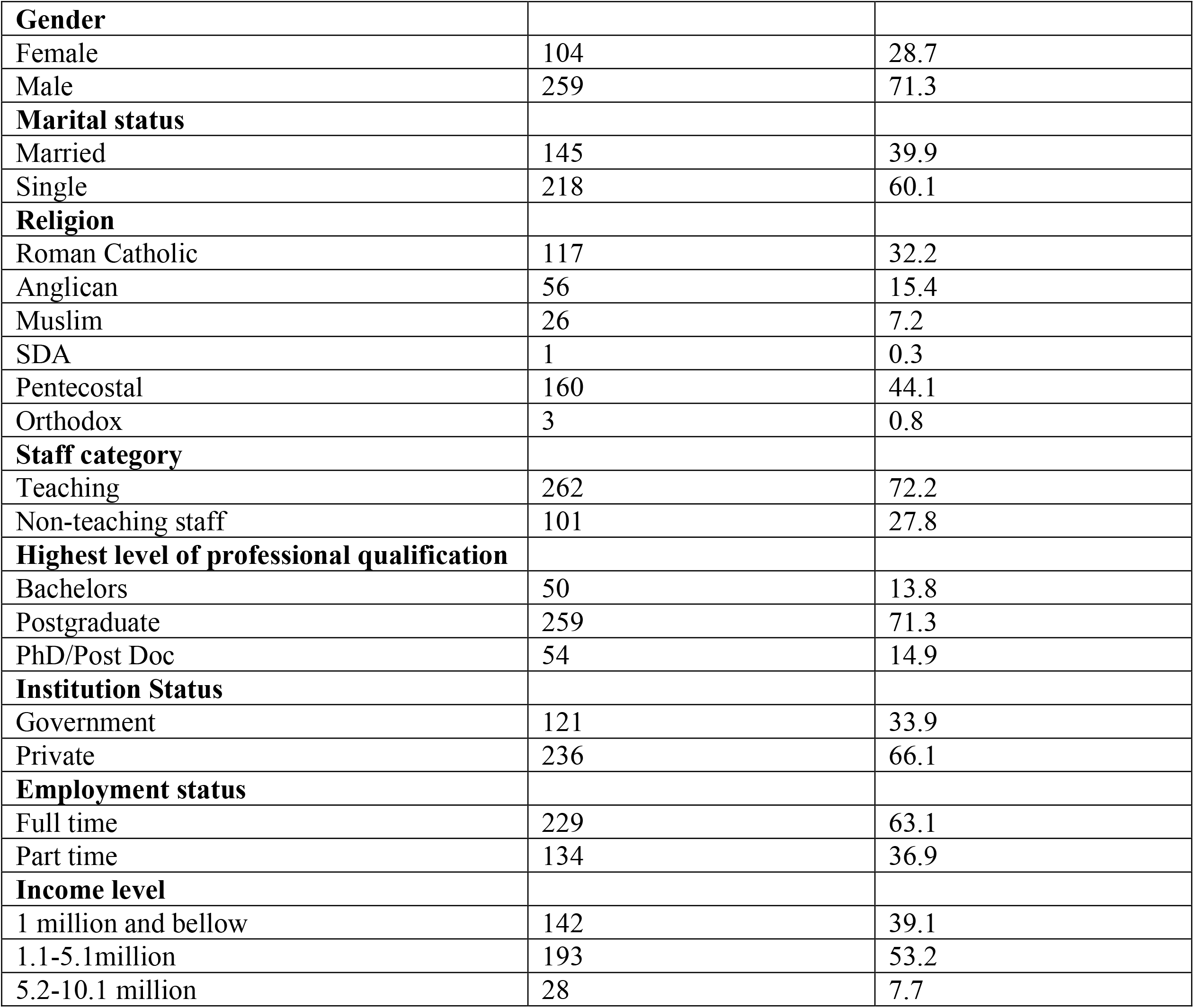
Demographic Characteristics of respondents.

### Intentions and willingness to receive COVID-19 Vaccines among teaching and non-teaching staff in higher institutions of learning in Uganda

Less than half 130(35.8%) of the respondents had the intention and willingness to receive COVID- 19 vaccine and more than half 233(64.2%) did not have the intention to receive COVID-19 vaccine (See **Figure *1***).

**Figure 1:**
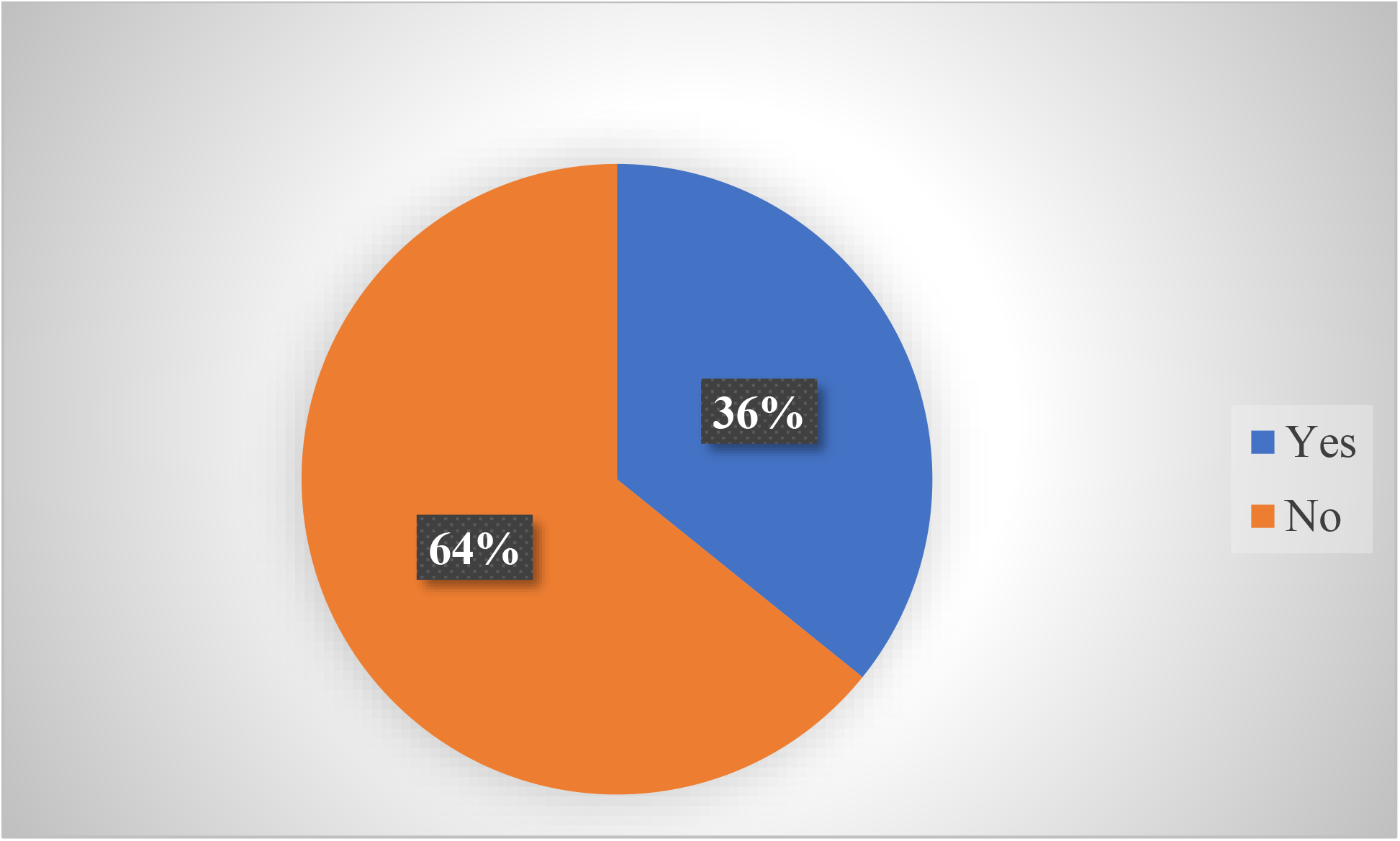
Intention and Willingness to receive COVID-19 vaccine among teaching and non-teaching staff in higher institutions of learning Factors influencing intentions and willingness to receive COVID-19 vaccine among teaching and non-teaching staff in selected institutions of higher learning in Kampala and Wakiso districts, Central Uganda

In our multivariate analysis, age between 30-39 (aOR=15491.54,95% CI=359-667551.9, P=0.000**, age between 40-49 (aOR=931, 95% CI=25.6-33816.4, p=0.000**), gender (aOR=4.66, 95% CI=2.06-1056, p=0.000*), staff category (aOR=7.71, 95% CI=4.34- 14.71,p=0.000**), ownership of the educational institution (aOR=0.08, 95% CI=0.032-0.206, p=0.000**), being under health insurance (aOR=200.62, 95% CI=29.6-1359.18, p=0.000*, residency (aOR=19.88, 95% CI=4.63-85.26, p=0.000*), knowing where to get COVID-19 vaccine (aOR=121.15, 95% CI=161.1-910.3, p=0.000**), COVID-19 vaccine is important for prevention of the infection (aOR=19.73, 95% CI=2.27-171.52, p=0.007*), minor side effects of COVID-19 vaccines are manageable (aOR=002, 95% CI= 0.00-0.15, p=0.000**), taking painkillers like paracetamol before receiving COVID-19 vaccine is not recommended (aOR=0.28, 95% CI=0.16- 0.495, p=0.000**), may take painkillers if you develop side effects after COVID-19 vaccine (aOR=8.67, 95% CI=4.87-15.43, p=0.000*), trust information of authorities on COVID-19 vaccine (aOR=0.03, 95% CI=0.002-0.387, p=0.008*), responsibility to protect others by getting vaccinated (aOR=0.36, 95% CI=0.21-0.62, p=0.000**), concerned about getting infected with COVID-19 from the vaccine (aOR=0.002, 95% CI=0.00-0.03, p=0.000*), trust the efficacy of the vaccine (aOR=0.04, 95%CI=0.02-0.11, p=0.000**), were statistically significantly associated with intentions and willingness to receive COVID-19 vaccine among teaching and non-teaching staff of higher institutions of learning (See **Table 3**).

### Level of knowledge on COVID-19 Vaccine

In this study, only 103 (28.4%) of the teaching and non-teaching staff in higher institutions of learning were more knowledgeable about COVID-19 vaccines and the majority 260(71.6%) were less knowledgeable about the vaccine (**See Figure 2**).

**Figure 2:**
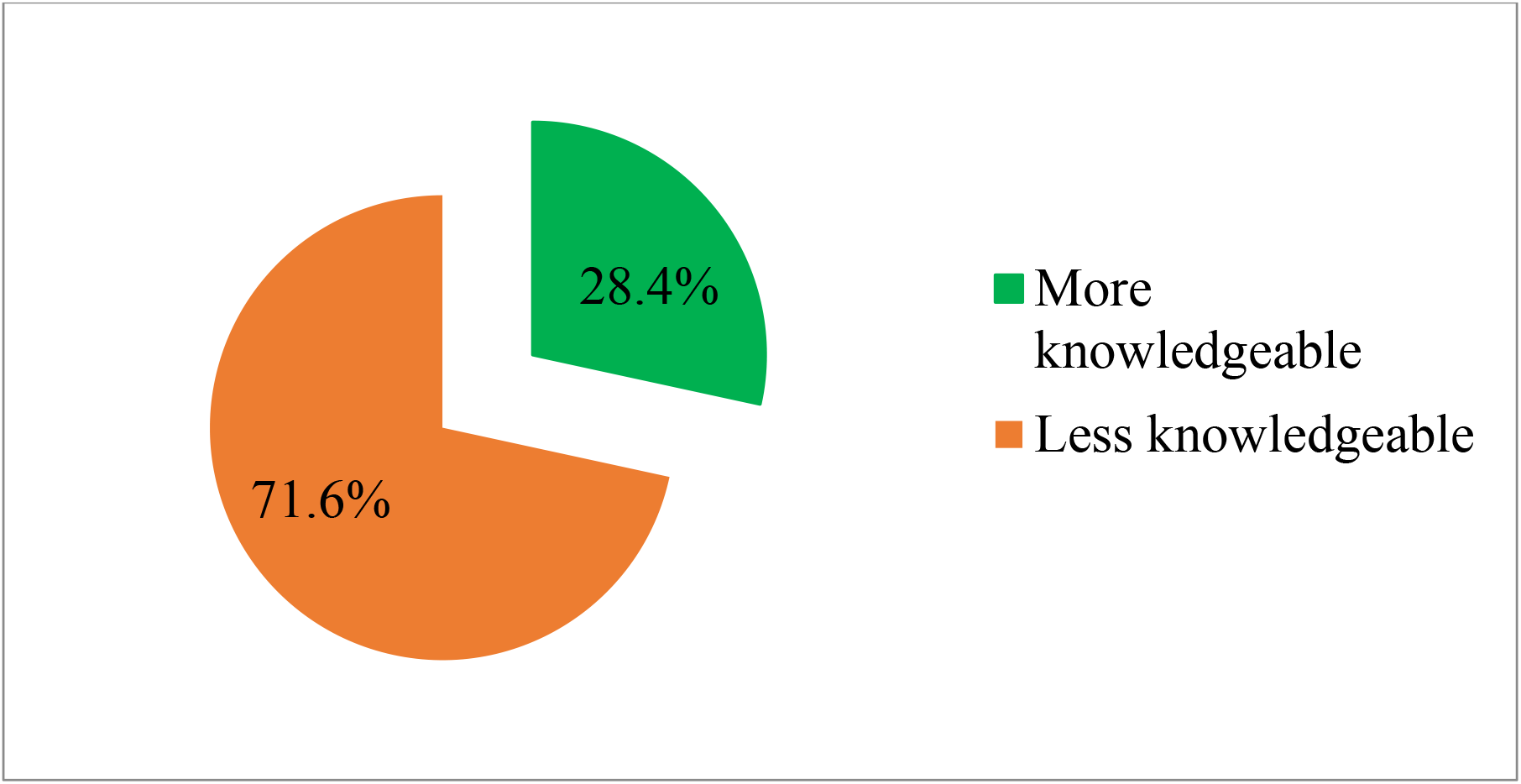
Level of knowledge on COVID-19 vaccine.

**Figure 3:**
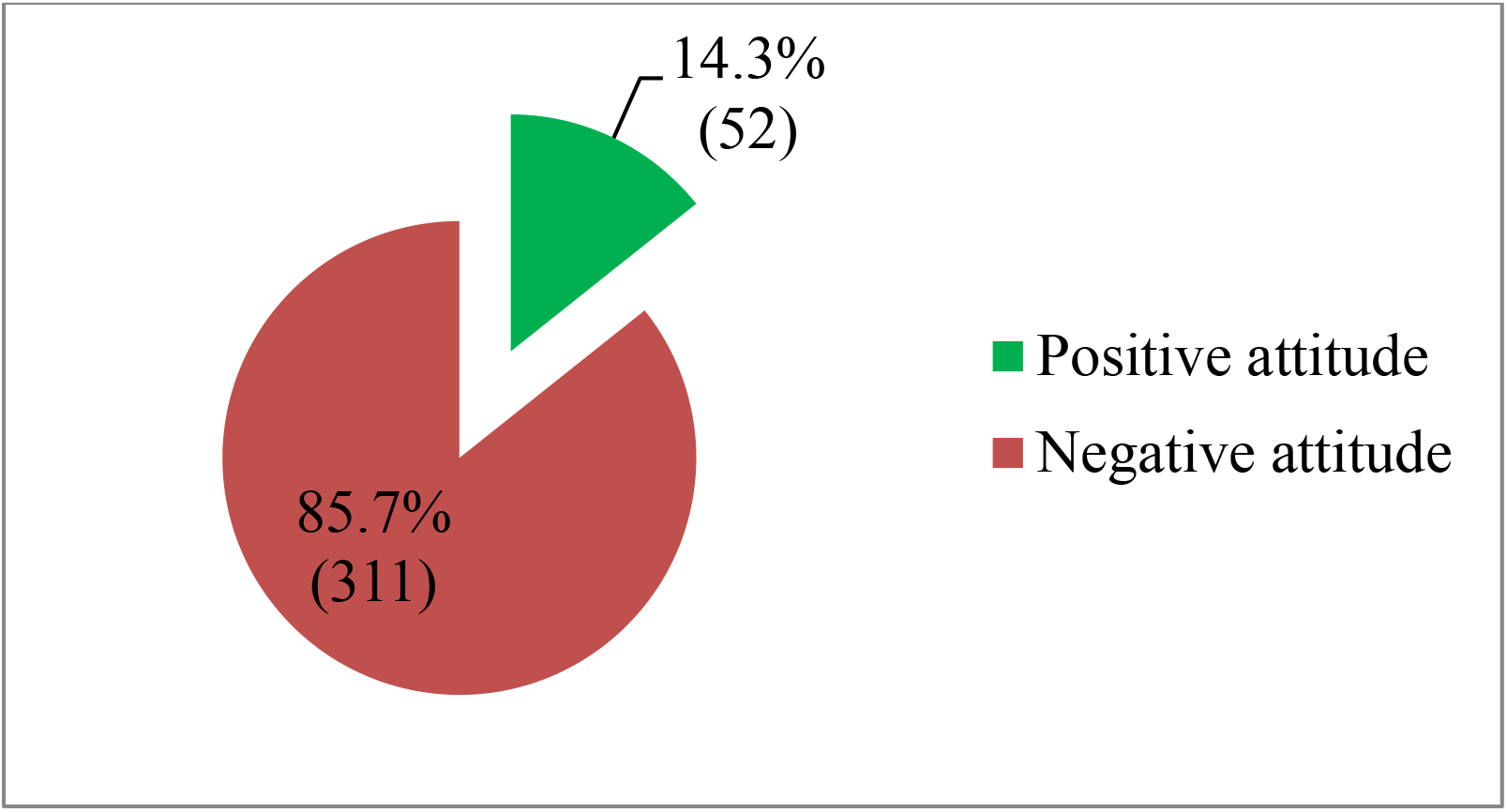
Attitude of respondents on intentions and willingness to receive COVID-19 vaccine.

### Attitude of respondents on intentions and willingness to receive COVID-19 vaccine

In this study, the majority 311(85.7%) of the teaching and non-teaching staff had negative attitude towards the intentions and willingness to receive COVID-19 vaccine, while only, 52(14.3%) had positive attitude towards intentions and willingness. The result further shows that, positive attitude significantly increases intentions and willingness to receive COVID-19 by 50 percent (OR=0.50, 95%CI: 0.28-0.91, p=0.023) (See **Table 2**).

**Table 2:** Attitude and knowledge of respondents on intentions and willingness to receive COVID-19 vaccine.

| Variable | Intention and Willingness to receive Covid-19 |  | Total | Unadjusted Odds ratio | p-value |
| --- | --- | --- | --- | --- | --- |
|  | Yes | No |  |  |  |
| **Level of knowledge on COVID-19** |  |  |  | Chi-square 5.315 | 0.021* |
| Positive attitude | 26(50) | 26(50) | 52 | 0.50(0.28-0.91) | 0.023* |
| Negative attitude | 104(33.4) | 207(66.6) | 311 | 1 |  |
| **Level of knowledge on COVID-19** |  |  |  | Chi-square 0.001(1) | 0.978 |
| More knowledgeable | 37(28.5) | 93(71.5) | 130 | 0.99(0.62-1.60) | 0.978 |
| Less knowledgeable | 66(28.3) | 167(71.7) | 233 | 1 |  |

However, respondents aged 20-29 (p=0.999) and 50-59 (aOR=33.5, 95% CI=0.17-6684.5, p=0.194), religious status (p=1.000), level of professional qualification (p=1.000), friends/workmate died of COVID-19 (aOR=1.36, 95% CI=0.21-8.63, p=1.000), COVID-19 vaccine is safe and effective in ending the pandemic (aOR=0.92, 95% CI=9.47-1.83, p=0.924), teachers should be prioritised for COVID-19 vaccine (aOR=0.66, 95% CI=0.38-1.14, p=0.132), not at risk of getting seriously ill of COVID-19 complications (p=0.994), simple and clear message on COVID-19 designed by the MoH (aOR=0.53, 95% CI=0.28-1.00, p=0.051), health workers have confidence in COVID1-9 vaccine (aOR=0.68, 95% CI=0.32-1.45, p=0.321) were not statistically significant (See ***Table 3***).

**Table 3:**
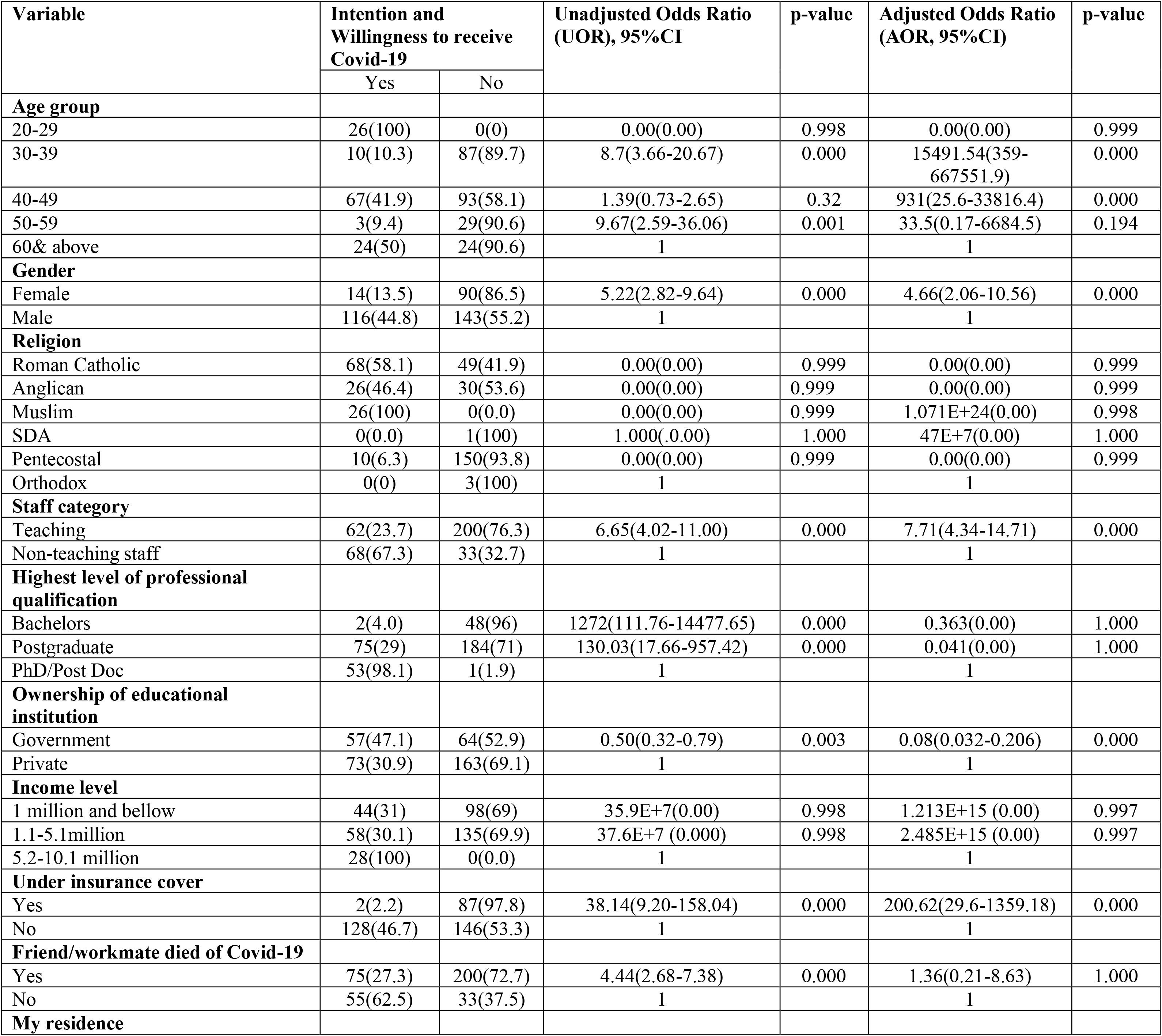
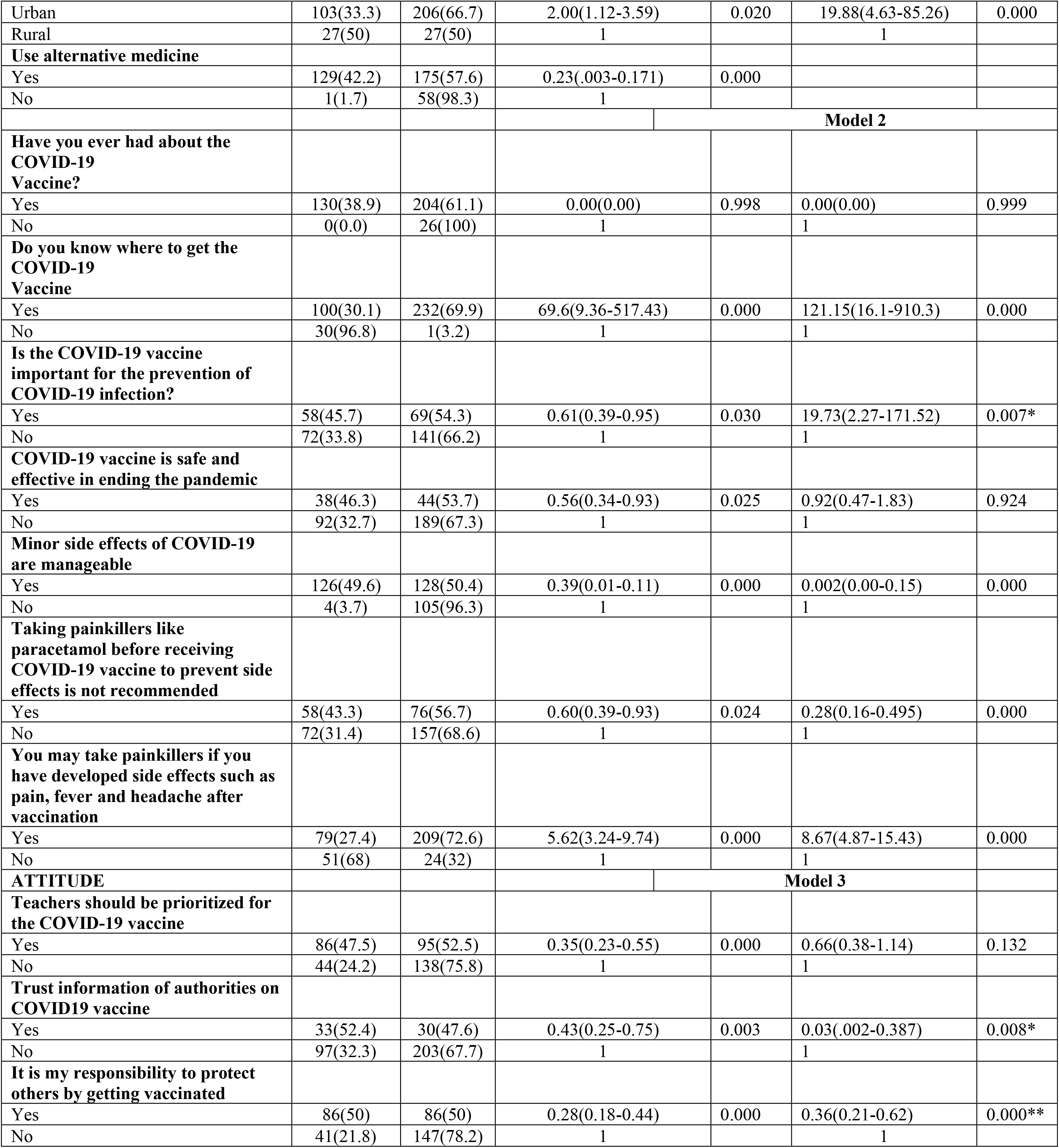
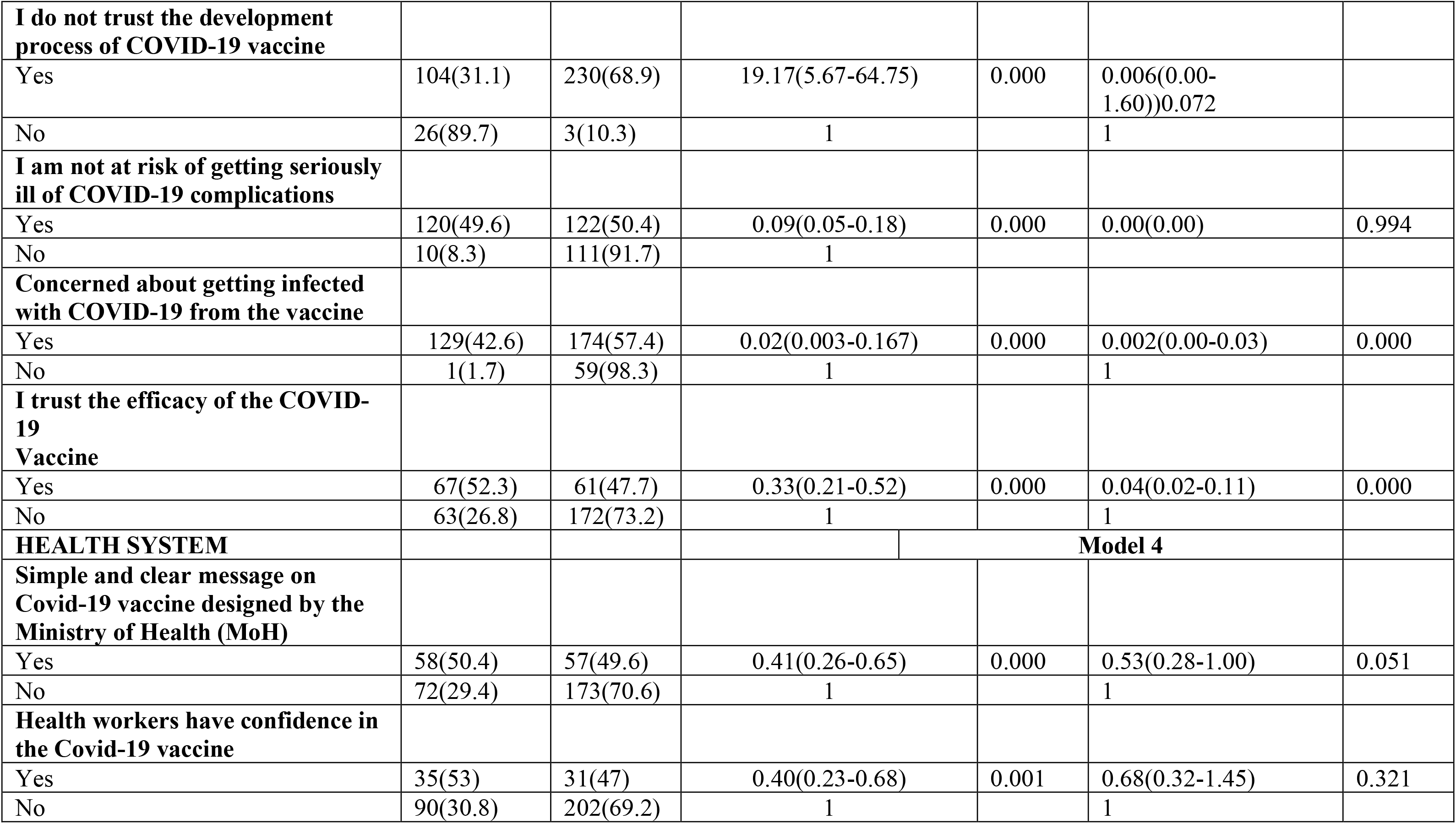
Factors influencing intentions and willingness to receive COVID-19 vaccine among teaching and non-teaching staff in selected higher institutions of learning in Kampala and Wakiso districts, Central Uganda.

In our univariate analysis, female teaching and non-teaching staff were more likely to have intentions and willingness to receive COVID-19 vaccine, compared to their male counterparts. Respondents aged 20-19 years of age were less likely to have intentions and willingness compared to teaching and non-teaching staff aged 60 years and above. In line with staff category, teaching staff were more likely to have intentions and willingness to receive COVID-19 vaccine compared to non-teaching staff and respondents with bachelor’s degree were more likely to have intentions and willingness to receive COVID-19 vaccine compared to respondents who had PhD/Post Doc. Moreover, teaching and non-teaching staff in government institutions were less likely to have intentions and willingness to receive COVID-19 vaccines compared to their counterparts in private higher institutions of learning and respondents whose friends or workmates died of the disease were more likely to have intentions and willingness to receive COVID-19 vaccine.

In terms of residency, teaching and non-teaching staff who reside within urban centres were more likely to have intentions and willingness to receive COVID-19 vaccine. Those who use alternative medicines were less likely to have intentions and willingness to receive COVID-19 vaccine. However, respondents who knew where to get COVID-19 vaccine were more likely to have intentions and willingness, respondents who said COVID-19 vaccine was important for prevention of the infection were less likely to have intentions and willingness to receive the vaccine, including teaching and non-teaching staff who said that the vaccines were safe and effective in ending the pandemic. Respondents who said that minor side effects of COVID-19 vaccine are manageable were less likely to have intentions and willingness.

In addition, teaching and non-teaching staff who said, take painkillers like paracetamol before receiving COVID-19 vaccines to prevent side effects were less likely to have intentions and willingness to receive COVID-19 vaccine, those who said, take painkillers when you have developed side effects such as pain, fever and headache after receiving the vaccine were more likely to have the intentions and willingness to receive COVID-19 vaccine. Respondents who said teachers should be prioritised for COVID-19 vaccines were less likely to intentions and willingness and those who said they trust information of the authorities on COVID-19 vaccine were less likely to have intentions and willingness, including those who said it was their responsibility to protect others by getting vaccinated.

Teaching and non-teaching staff who said they do not trust the development process of COVID- 19 vaccine were more likely to have intentions and willingness to receive COVID-19 vaccine compared to who said they trust the process. On the contrary, respondents who said they were not at risk of getting seriously ill of COVID-19 complications were less likely to have intentions and willingness compared to those who said they were not at risk of COVID-19 complications.

Also, some respondents were concerned of getting infected with COVID-19 from the vaccines, these were less likely to have intentions and willingness. Those who trust the efficacy of the vaccines were less likely to have intentions and willingness. In line with the health system, respondents who said there was simple and clear message on COVID-19 vaccine designed by the Ministry of Health were less likely to have intentions and willingness to receive the vaccine and those who said health workers have confidence in the COVID-19 vaccine were as well less likely to have intentions and willingness to receive the vaccine (See **Table *3***).

### Communication channels from which respondents ever got messages on COVID-19 vaccine

Our study found out that, some of the common channels of communication from which respondents ever got messages on COVID-19 vaccine included, radio 52(9.2%), social media 130(23.1%), television 278(49.4%), newspapers 26(4.6%), and Ministry of Health website 77(13.7%) (**See Figure *4***).

**Figure 4:**
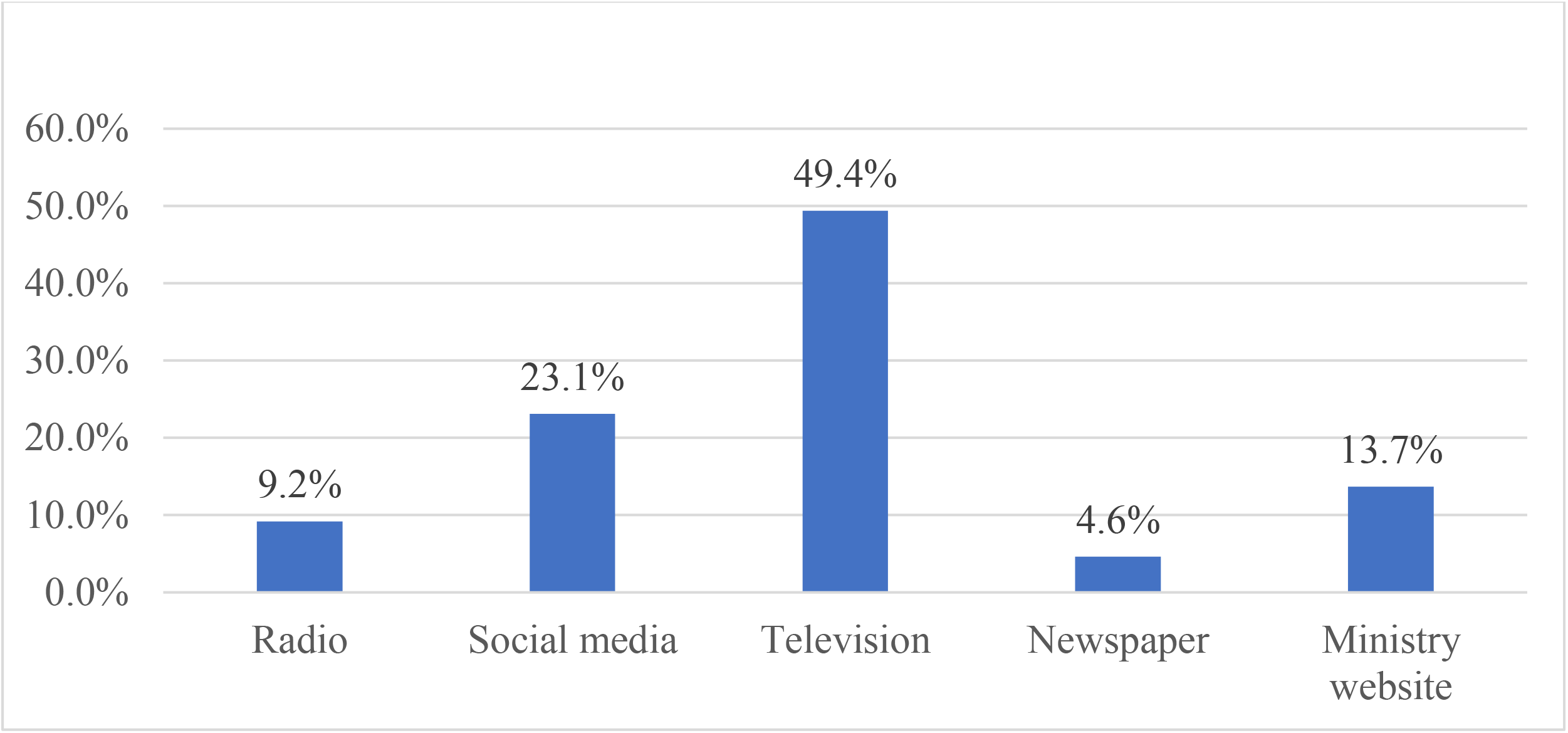
Communication channels from which respondents ever got messages on COVID- 19 vaccine.

## Discussion

### Intentions and willingness to receive COVID-19 Vaccines among teaching and non-teaching staff in higher institutions of learning in Uganda

In this study, there was low 130(35.8%) intention and willingness to receive COVID-19 vaccine, this was similar to a study conducted in Ethiopia among residents which showed that, less than one third of the participants were willing to accept the vaccines [30], similarly, 33.7% intention was recorded among HIV positive patients in Ethiopia [31]. Contrary to the findings of this current study, according to a study conducted in Macao, more than half (62.3%) of the participants had intention to receive COVID-19 vaccine [32]. Similarly, more than half (55%) adults with chronic conditions in Ethiopia had the willingness to receive COVID-19 vaccine [33] and 54.5% had intentions to receive COVID-19 vaccine among primary and secondary teachers in North-Western Ethiopia [34]

A study conducted in the United States of America noted that, 57.6% of the adults intended to receive COVID-19 vaccines, about 1 in 10 adults did not intend to receive the vaccine and 3 in 10 individuals were not sure if they should receive the COVID-19 vaccine [35], this percentage was lower than a study conducted by Reiter in the USA, which found 69% willingness to receive the vaccines [36]. A low percentage of willingness to receive COVID-19 vaccine appears to be less likely in achieving herd immunity as based on basic reprotection number estimates [37] Moreover, a study conducted among Canadian adults showed that 79.8% intentions to receive COVID-19 vaccine [38], the willingness and intention to receive COVID-19 vaccine in Canada was more pronounced compared to that of USA. Willingness to receive the vaccine in the UK was 76.9% which was slightly similar to that of Canada [39], and a survey conducted among European countries indicated that intentions and willingness to receive COVID-19 vaccine ranged between 62% to 80% [40]. Among the Lebanese adults, 40% strongly disagree with receiving the COVID- 19 vaccine [41].

Respondents aged 30-39 and 40-49 years were more likely to have intentions and willingness to receive COVID-19 vaccine and there was statistically significant relationship between age and willingness to receive COVID-19 vaccine compare to teaching and non-teaching staff who were 60 years and above. This current study is contrary to a study conducted in Canada to ascertain intentions to receive COVID-19 vaccine which showed that, individuals older than sixty years were more likely to have intentions to receive the vaccine compared to those in the younger age group [38].

In terms of gender, our study found that females were more likely to have intentions and willingness to receive COVID-19 vaccine compared to their male counterparts and there was statistically significant relationship between gender and intentions and willingness to receive COVID-19 vaccine. Contrary to our study, a study among Lebanese adults showed that intentions and willingness to receive the COVID-19 vaccine was low among females [41], a study conducted by Kaadan *et al* also noted higher intentions among males compared to females [42]. More studies in different parts of the world confirmed acceptance of the vaccine among males was higher [43, 44, 45]. These gender differences in COVID-19 vaccine acceptance may be associated with the psychological differences between females and males which indicated that anxiety and depression are higher in women compared to men [46, 47]. Also, females had higher fear of injections and side effect concerns [48]. Similarly, willingness to receive COVID-19 vaccines among teachers and students in Qatar was higher among males compared to their female counterparts [49]. Vaccine acceptance was low among females according to a study conducted by Bono *et al* [50]. However, another study conducted in Uganda by Muhindo *et al*., on COVID-19 vaccine acceptance was higher among women compare to men [51]. Also, a global population study by Huang *et al.,* showed that, females were more likely to have intentions to receive COVID-19 [52].

In line with staff category, teaching staff were more likely to have intentions and willingness to receive COVID-19 vaccine compared to non-teaching staff and respondents with bachelor’s degree were more likely to have intentions and willingness to receive COVID-19 vaccine compared to respondents who had PhD/Post Doc. Contrary to our study, a study conducted in Congo indicated that level of education was associated with acceptance of COVID-19, the higher the education level, the higher the acceptance [53], similarly in the United States, those with a lower level of education had disparity in willingness to receive COVID-19 vaccine [54]. teachers and students with or perusing postgraduate degrees were more willing to receive vaccines compared to those with diplomas or bachelor’s degrees [49]. Moreover, other studies have indicated education as one of the key factors influencing intentions and willingness to receive the COVID-19 vaccine [55, 56]. A study conducted in Ethiopia among adults in the general population showed that, higher education levels from secondary education and above were associated with willingness to receive the vaccine [57].

Moreover, teaching and non-teaching staff in government institutions were less likely to have intentions and willingness to receive COVID-19 vaccines compared to their counterparts in private higher institutions of learning, this could be related to the fact that many private institutions had put more strict measures for every staff member to receive the COVID-19 vaccine, contrary to our study, a study conducted in Ethiopia showed that participants who were not employed by the government were less likely to have intentions to receive COVID-19 vaccine [58] and respondents whose friends or workmates died of the disease were more likely to have intentions and willingness to receive COVID-19 vaccine. Similarly, a study conducted among the elderly in Italy showed that respondents whose relatives or family member had died of COVID-19 were more likely to have willingness to receive vaccine [59] and a study conducted by Elhadi *et al* among health workers showed that COVID-19 related death of a friend or family member was negatively associated with willingness and intentions to receive COVI-19 vaccine while having a friend or family member infected with COVID-19 increased the likelihood of willingness and intention to receive the vaccine [60].

This current study also found out that, staff members who are under health insurance were more likely to have intentions and willingness to receive COVID-19 vaccine compare to those who are not covered by health insurance, similarly, a global population study on intentions and willingness to receive COVID-19 vaccine showed that participants who were under insurance cover were more likely to have intentions to receive the vaccine [52].

Respondents who knew where to get COVID-19 vaccine were more likely to have intentions and willingness to receive COVID-19 vaccine and there was statistically significant relationship, also, teaching and non-teaching staff members who knew that COVID-19 vaccine was important for prevention of the infection were more likely to have intention and willingness. Knowing where to get the vaccines from would probably have influence intentions and willingness, in addition with having knowledge about the vaccine, a study conducted in the USA showed that limited knowledge on COVID-19 vaccines contributed to increased hesitancy in intentions and willingness to receive the vaccines [61].

Moreover, respondents who said that minor side effects of COVID-19 vaccines are manageable were less likely to have intentions and willingness to receive COVID-19 vaccine and there was statistically significant relationship. Similarly, respondents who said taking painkillers like paracetamol before receiving COVID-19 vaccine is not recommended were less likely to have intentions and willingness to receive COVID-19, whereas, teaching and non-teaching staff who said one may take painkillers if you develop side effects after COVID-19 vaccine were more likely to have intentions and willingness to receive the vaccine and there was a statistically significant relationship. Similarly, Elhadi et al noted that, having concerns about the side effects of the COVID-19 vaccine contributed to vaccine hesitancy and reduced the willingness to receive the vaccine [60]. It therefore very important to improve the perception and knowledge of teaching and non-teaching staff of higher institutions of learning in order to improve their intentions and willingness to receive the vaccines.

In this study, some respondents said they trust information of authorities on COVID-19 vaccine, although those who trusted the information of authorities on COVID-19 did not translate to having intentions and willingness to receive the vaccine. Similarly, respondents who said it was their responsibility to protect others by getting vaccinated were less likely to have intentions and willingness to receive the vaccine. A study conducted by Lang *et al* noted that, individuals’ trust in sources of message from health authorities or the ministry of health had great potential for COVID-19 vaccine uptake among the general population [62]. Vaccine hesitancy has been recorded as the 10^th^ threat to global health by the WHO [63] and among the 10 global health issues to handle in 2021 [64]. This implies that COVID-19 vaccine issues should be reflected in all health policy statements of the MoH and should be handled as one of the priorities of the health system.

Respondents who had concern about getting infected with COVID-19 from the vaccine and those who said they trusted the efficacy of the vaccines were less likely to receive to have intentions and willingness to receive COVD-19 vaccine and there was statistically significantly association. Similarly, Elhadi et al noted that having concerns about the side effects of the COVID-19 vaccine contributed to vaccine hesitancy and reduced the willingness to receive the vaccine [60]. Moreover, some individuals got COVID-19 vaccines because of perceived social responsibility, having a desire to get back to normal work and fear of getting a serious illness or serious complications [65].Gidengil *et al* noted that if H1N1 vaccine availability changed people’s actual vaccination intention [66], we would note whether the intentions of teachers in higher institutions of learning would be different from when COVID-19 vaccines are available and have been prioritised by the government to receive the vaccine. moreover, some individuals do not believe in the efficacy and effectiveness of vaccines against viral infections like COVID-19, especially those who had ever received flu vaccines and developed multiple upper respiratory tract infections during the year they took the vaccine [67].

In this study only 103 (28.4%) of the teaching and non-teaching staff in higher institutions of learning were more knowledgeable about COVID-19 vaccines and respondents whose friends/workmate died of COVID-19 were more likely to have intentions and willingness to receive COVID-19 vaccine, moreover, teaching and non-teaching staff who said COVID-19 vaccine is safe and effective in ending the pandemic were less likely to have intentions and willingness. Our study findings were contrary to a study conducted in Ethiopia found that, COVID- 19 vaccine knowledge among the general population was 74% [57]. This is lower than knowledge about vaccines in England which is at 83% and France at 81.2%, and the finding on COVID-19 vaccine knowledge was found to be higher than that of Bangladesh at 62.1% and West India at 35.5% [68, 69]. In addition, a study conducted in the USA showed that limited knowledge and mistrust in the COVID-19 vaccines contributed to increased hesitancy in intentions and willingness to receive the vaccines [61].

Also, respondents who said teachers should be prioritised for COVID-19 vaccine, they are not at risk of getting seriously ill of COVID-19 complications, simple and clear message on COVID-19 designed by the Ministry of Health and health workers have confidence in the COVID1-9 vaccine were less likely to have intentions and willingness to receive the vaccine. Similarly, the WHO recommends that, teachers should be considered important in the prioritization process for COVID-19 vaccination, during a period of community transmission, teachers are included in stages II in a situation where vaccine availability is between 11–20% and III when vaccine availability is 21–50% [70], moreover, UNESCO and Education International also call for teachers to be prioritized to receive the COVID-19 vaccine once older and other high-risk populations, as reopening of educational institutions safely will keep them open as long as possible [27].COVID-19 vaccine should be prioritized for teachers according to UNICEF [71].

Benefits of vaccinating teachers and other professionals working in these education institutions include: ensuring continuity of teaching in-person, reducing teachers’ likelihood of infection, improves student safety; and increasing confidence in parents that education educations are safe places to be in [27, 28]. Teaching and non-teaching staff who said they do not trust the development process of COVID-19 vaccine were more likely to have intentions and willingness to receive COVID-19 vaccine compared to who said they trust the process. On the contrary, respondents who said they were not at risk of getting seriously ill of COVID-19 complications were less likely to have intentions and willingness compared to those who said they were not at risk of COVID-19 complications. Similarly, a study conducted in Bangladesh on COVID-19 vaccine acceptance showed that perceived severity of the infection influenced one’s willingness to accept the vaccination [72].

Also, some respondents were concerned of getting infected with COVID-19 from the vaccines, these were less likely to have intentions and willingness to receive COVID-19 vaccine. Those who trust the efficacy of the vaccines of less likely to have intentions and willingness and in line with the health system, respondents who said there was simple and clear message on COVID-19 designed by the Ministry of Health were less likely to have intentions and willingness to receive COVID-19 vaccine and those who said health workers have confidence in the COVID-19 vaccine were as well less likely to have intentions and willingness to receive COVID-19 vaccine. Similarly, more studies conducted in different parts of the world on COVID-19 vaccination indicated that, some of the key factors influencing vaccine acceptance were attitude, belief and safety concerns about the COVID-19 vaccine [73, 74, 75, 76]. Some of the reasons for vaccine acceptance included a feeling of collective responsibility and being part of the fight against the virus [77], this may be related to psychological and motivational determinants as a sense of responsibility among individuals for population health and social solidarity [78].

### Study Strength and Limitations

Our study had several strengths, firstly, the large sample size, secondly this was the first study in Uganda which was done among teaching and non-teaching staff of higher institutions of learning on intentions and willingness to receive COVID-19 vaccines, thirdly, we maintained confidentiality and ensured correctness and consistency of the recorded data. Fourth, our research tools were fully pretested and checked for accuracy and correctness, only fully filled questionnaire were included for analysis. However, the study could not be used to analyse intentions and willingness of teaching and non-teaching staff over time as it was a snapshot, limitations of confounding bias was controlled by restricting participation to only those who had not yet been vaccinated at the time of the study and had worked within the selected institutions of higher learning within Kampala and Wakiso for at least six months so as to arrive at conclusive results.

## Conclusion

Intentions and willingness to receive COVID-19 vaccine was very low. This was attributed to age gender, staff category, ownership of the educational, being under health insurance, residency, knowledge on COVID-19 vaccine and where to get the vaccine, trust information of authorities and vaccine efficacy, concern about getting infected with COVID-19 from the vaccine. Efforts should be directed to promote the effectiveness of the vaccines among teaching and non-teaching staff of higher institutions of learning, especially in government institutions to increase the willingness.

## Data Availability

All relevant data are within the manuscript and its Supporting Information files.

## Abbreviations

ACT: Access to COVID-19 Tools
aOR: Adjusted Odds Ration
CI: Confidence Interval
MoH: Ministry of Health
uOR: Unadjusted Odds Ration
UNESCO: United Nations Educational, Scientific and Cultural Organization
UK: United Kingdom
UNICIEF: The United Nations International Children’s Emergency Fund
WHO: World Health Organization

## Acknowledgement

We thank Clarke International University Research Ethics Committee (CIUREC) for granting us ethical approval for the study. We appreciate all our research assistants for the support they rendered during the data collection process.

## Consent for Publication

No application

